# Exploring the Role of Oxidative Stress in Metallic Taste During Head and Neck Cancer Treatment: A Study of Salivary Malondialdehyde and Therapeutic Interventions

**DOI:** 10.1101/2025.04.28.25326099

**Authors:** Guillaume Buiret, Thierry Thomas-Danguin, Gilles Feron

## Abstract

**Introduction:** Head and neck cancers (HNC) treatments often cause a metallic taste (MT), adversely affecting patients’ quality of life. This study aims to investigate the lipoperoxidation hypothesis of MT by examining salivary malondialdehyde (MDA) levels, a marker of oxidative stress, in HNC patients undergoing treatment.

**Methods:** This prospective cohort study included 44 newly diagnosed HNC patients. Saliva samples were collected before, during and up to one year after the HNC treatment. Analyses including MDA and other markers were performed. Additionally, a bovine lactoferrin mouthwash was evaluated for its efficacy in alleviating MT.

**Results:** Out of the 44 patients, 12 (27.2%) reported MT, primarily during treatment phases. Salivary MDA levels significantly increased during radiotherapy, peaking mid-treatment, before declining post-treatment. Despite this fluctuation, no significant relationship was found between MDA levels and MT. Bovine lactoferrin mouthwash alleviated MT at least partially in 63.2% of the occurrences. Other salivary markers such as protein concentration, antioxidant properties, catalase activity, and superoxide dismutase activity showed no significant link to MT.

**Discussion:** The increase in MDA levels during radiotherapy indicated heightened oxidative stress. However, the lack of a significant association between MDA and MT suggests other factors may contribute to MT development. The partial efficacy of lactoferrin mouthwash highlighted a potential benefits. Future research should explore other mechanisms, such as the role of oral microbiota, to better understand and manage MT in HNC patients.

## Introduction

Head and neck cancers (HNC) include cancers of the oral cavity, pharynx, and larynx, with tobacco use, alcohol consumption, and human papillomavirus (HPV) being the primary risk factors. In France they are the fifth most common group of cancers (1). The treatments are multimodal, including surgery, radiotherapy, chemotherapy targeted therapies and, more recently, immunotherapy. Due to the cancer end/or its treatments, one of the lesser-studied but clinically significant symptom is the development of a metallic taste (MT). This altered taste perception can significantly impact patients’ quality of life, affecting their nutritional intake and overall well-being (2). However, little or nothing is known about the origin of MT in HNC. There are two main hypotheses (3): the release of inhibition of the facial nerve on the glossopharyngeal nerve (4, 5) and the oral mucosa lipoperoxidation (6). Malondialdehyde (MDA) is the final stable product of lipid peroxidation and is considered a reliable marker of oxidative stress. It has been documented in various inflammatory or precancer diseases (7-9), HNC (8) as well as in smokers (10). Given that saliva plays also a crucial role in oral health and taste perception (11, 12), it is also a relevant medium for assessing oxidative stress marker like MDA (7, 10, 13, 14).

Bovine lactoferrin was shown to alleviate MT induced by ferrous sulfate mouthwash in healthy volunteers (6). It has immunoregulatory mechanisms, especially in vitro and in animals supplemented per os (15). But no trial has yet been performed to evaluate its putative effect on MT in a context of gustatory trouble.

By conducting serial saliva collections and analyses over a one-year period on patients treated for a HNC, we sought to determine if changes in salivary MDA concentrations or other salivary composition parameters were associated with the onset and severity of MT, thereby providing insights into potential markers and underlying mechanisms of MT in HNC patients. We also used bovine lactoferrin mouthwash to try to alleviate MT.

## Methods

### Study Design

This prospective cohort study was conducted over a two-year period, involving 44 patients newly diagnosed with HNC. Participants were recruited from the ENT departments of a regional teaching hospital. They were followed up for one year after the end of their treatment, unless a recurrence or another cancer occurred. At the end of the study, they were categorized into two distinct groups based on the presence or absence of reported MT.

### Participants

Inclusion criteria were: adults aged 18 years and older, newly diagnosed with HNC, and scheduled to undergo treatment (surgery, radiation, and/or chemotherapy). Exclusion criteria included previous treatment for HNC, other malignancies, having a total laryngectomy, or being pregnant.

### Data Collection

The population’s characteristics were assessed at the beginning of the study. Assessments were performed on each patient at regular intervals: before any treatment; after surgery if any; midway and upon completion of radio(chemo)therapy if any; and at 3-6-9 and 12 months.

Several questionnaires were used: EORTC QLQ30 (9) and HN35 (10) questionnaires to evaluate Quality of Life (QoL), a specific questionnaire to assess MT impact (11) in patients complaining about MT, measurement of weight and a Visual Analog Scale (VAS) to quantify food intake (12) ranging from 0 (total abstention) to 10 (normal eating).

An oral examination looking for mucositis and/or candidiasis was performed at each assessment.

Similarly, unstimulated whole saliva samples were also collected. Participants were instructed to refrain from eating, drinking, smoking, or performing oral hygiene activities for at least one hour before sample collection. Saliva was collected by having participants drool into sterile tubes over a ten-minute period. Saliva flow was calculated from the weight of saliva assuming that 1 g is equal to 1 mL. Samples were immediately placed on ice, transported to the laboratory, weighed and stored at - 80°C until analysis. Aliquotes of saliva samples were previously defrosted and then centrifuged 15 min at 15 000 g. Analyses were performed on the resulting supernatants.

### Salivary MDA analysis

To summarize, salivary MDA levels were measured using a modified thiobarbituric acid reactive substances (TBARS) assay. 2,6-di-tert-butyl-4-méthylphénol (BHT) was added to an aliquot of whole saliva samples until a concentration of 280µM and then the saliva samples were clarified by centrifugation 15 min at 15000 g at 4°C. 150µL of supernatant were mixed with thiobarbituric acid in duplicates and incubated at 60°C for 30 minutes. The fluorescence of the reaction mixture was measured (excitation: 532 nm; emission : 553 nm) using a microplate reader (Ensight, PerkinElmer, Waltham, MA). MDA concentrations were calculated using the linear regression obtained from a standard range prepared with known concentrations of MDA. The salivary MDA concentration is the mean of the 2 replicates expressed in µM MDA equivalent.

### Other biochemical analysis

If appropriate, all enzyme activities were expressed in milli International Enzyme Activity Units (mIU) per ml of saliva. One IU was defined as the amount of enzyme that catalyses the conversion of 1 µmol of substrate per minute. Protein concentration (mg/mL) was measured with a Quick Start Bradford protein assay (Bio-Rad, France) (excitation: 595 nm) using a microplate reader (Ensight, PerkinElmer, Waltham, MA).

### Total antioxidant status quantification

Total antioxidant status was determined using an ORAC Assay kit (Cell Biolabs Inc, Oxiselect™, San Diego, CA). This assay uses the free radicals initiator AAPH (2,2⍰-azobis-2-methyl-propanimidamide dihydrochloride). Peroxyl radicals then oxidize fluorescein with a consequent loss of fluorescence. Trolox (6-hydroxy-2,5,7,8-tetramethylchroman-2-carboxylic acid), a water-soluble vitamin E analogue, served as a standard to inhibit fluorescein decay in a dose dependent manner. Kinetics of the intensity of fluorescence was recorded at 37°C (excitation: 485 nm; emission : 538 nm) using a microplate reader (Ensight, PerkinElmer, Waltham, MA). The antioxidant capacity of saliva was expressed as micromolar Trolox equivalents per mL.

### Catalase

Catalase Activity was determined using a Catalase Fluorescent Activity Kit (Arbor Assay, Ann Arbor, MI, USA) by means of a fluorescent reaction coupled to HRP (horseradish peroxidase) which permit following the rate of oxidation of hydrogen peroxide added to samples. Increasing levels of catalase in the samples causes a decrease in hydrogen peroxide concentration and a reduction in fluorescent product (excitation at 570 nm, emission at 590nm) determined using a microplate reader (Ensight, PerkinElmer, Waltham, MA). A bovine catalase standard is used to generate a standard curve. Results were expressed in terms of units of catalase activity per mL of saliva.

### SOD

SOD Activity was determined by SOD Assay Kit (Sigma-Aldrich, Saint-Louis, MI) using Dojindo’s highly water-soluble tetrazolium salt, WST-1 (2-(4-Iodophenyl)-3-(4-nitrophenyl)-5-(2,4-disulfophenyl)-2Htetrazolium, monosodium salt), producing a water-soluble formazan dye upon reduction with a superoxide anion. The rate of the reduction with O_2_ are linearly related to the xanthine oxidase (XO) activity, and is inhibited by SOD. The IC50 (50% inhibition activity of SOD) is determined by absorbance reading at 440nm using a microplate reader (Ensight, PerkinElmer, Waltham, MA). Since the absorbance at 440 nm is proportional to the amount of superoxide anion, the SOD activity as an inhibition activity could be quantified by measuring the decrease in the color development. A bovine SOD standard (Sigma-Aldrich, Saint-Louis, MI) was used to generate a standard curve. Results were expressed in terms of units of SOD activity per mL of saliva.

### GST

Glutathion S Transferase Pi (GSTp) was quantified using Enzyme-Linked Immunosorbent Assay kit from Abbexa Ltd. (Cambridge, UK). The concentrations were obtained in ng/mL of saliva.

### Bovine lactoferrin mouthwash

A mouthwash of 10ml with a lactoferrin-concentration of 50mg/l was proposed to patients who complained of a spontaneous MT. Lactoferrin powder, produced by Ingredia, France, was diluted with Evian water.

### Statistical Analysis

Descriptive statistics were used to summarize the demographic and clinical characteristics of the study population. Quantitative data were compared using mean comparisons through ANOVA, while qualitative data were analyzed using the chi-square test. The primary outcome was the association between salivary MDA levels and the presence of MT. Mean comparison (with t-tests if the distribution was normal or Wilcoxon test if not) of salivary MDA according to the presence or not of MT was used to assess the relationship between MDA levels and MT.

#### Ethical Considerations

This study was performed in line with the principles of the Declaration of Helsinki and adhered strictly to the French ethical guidelines and principles, with all participating patients providing informed consent prior to their involvement. Approval for the study was granted by the CPP Est I, Dijon, France (#2017-A03641-52). Additionally, the study is registered on ClinicalTrials.gov (NCT03558789).

## Results

### Participant Characteristics

A total of 44 patients were enrolled in the study. The flowchart of the study is presented in Figure 1. Table 1 displays the demographic characteristics of the study population at inclusion according to the MT status. Patients were predominantly male, of middle age, and with advanced Tumor and Node stage. The treatment modalities were predominantly multimodal.

**Table 1:** studied population characteristics at inclusion according to metallic taste status. MT: metallic taste

|  | MT | No MT | p-value |
| --- | --- | --- | --- |
| Sex: males n(%) ; females n(%) | 9 (75%), 3 (25%) | 25 (78.1%), 7 (21.9%) | 1 |
| Mean age at diagnosis ± SD (year) | 61.7 ± 10.2 | 63.6 ± 9.8 | 0.58 |
| Location n (%):<br>Hypopharynx<br>Larynx<br>Oropharynx<br>Oral Cavity<br>Unknown primary | 0 (0%)<br>1 (8.3%)<br>4 (33.3%)<br>5 (41.7%)<br>2 (16.7%) | 3 (9.4%)<br>7 (21.9%)<br>10 (31.2%)<br>9 (28.1%)<br>3 (9.4%) | 0.57 |
| T stage n (%):<br>1<br>2<br>3<br>4<br>x | 3 (25%)<br>6 (50%)<br>1 (8.3%)<br>0 (0%)<br>2 (16.7%) | 6 (18.7%)<br>12 (37.5%)<br>7 (21.9%)<br>4 (12.5%)<br>3 (8.8%) | 0.69 |
| N stage n (%):<br>0<br>1<br>2a<br>2b<br>2c<br>3a | 4 (33.3%)<br>4 (33.3%)<br>1 (8.3%)<br>2 (16.7%)<br>0 (0%)<br>1 (8.3%) | 16 (50%)<br>6 (18.7%)<br>3 (9.4%)<br>3 (9.4%)<br>2 (6.2%)<br>2 (6.2%) | 0.77 |
| Smoking at inclusion n (%):<br>Yes<br>No<br>Quit | 3 (25%)<br>3 (25%)<br>6 (50%) | 17 (50%)<br>2 (6.2%)<br>13 (40.6%) | 0.11 |
| Treatment<br>Withdrawal of consent before treatment<br>Chemotherapy, surgery, radiotherapy<br>Surgery<br>Surgery, radiotherapy<br>Surgery, radiochemotherapy | 0 (0%)<br>0 (0%)<br>2 (16.7%)<br>7 (58.3%)<br>3 (25%) | 5 (15.6%)<br>1 (3.1%)<br>12 (37.5%)<br>5 (15.6%)<br>9 (28.1%) | 0.54 |

**Figure 1.**
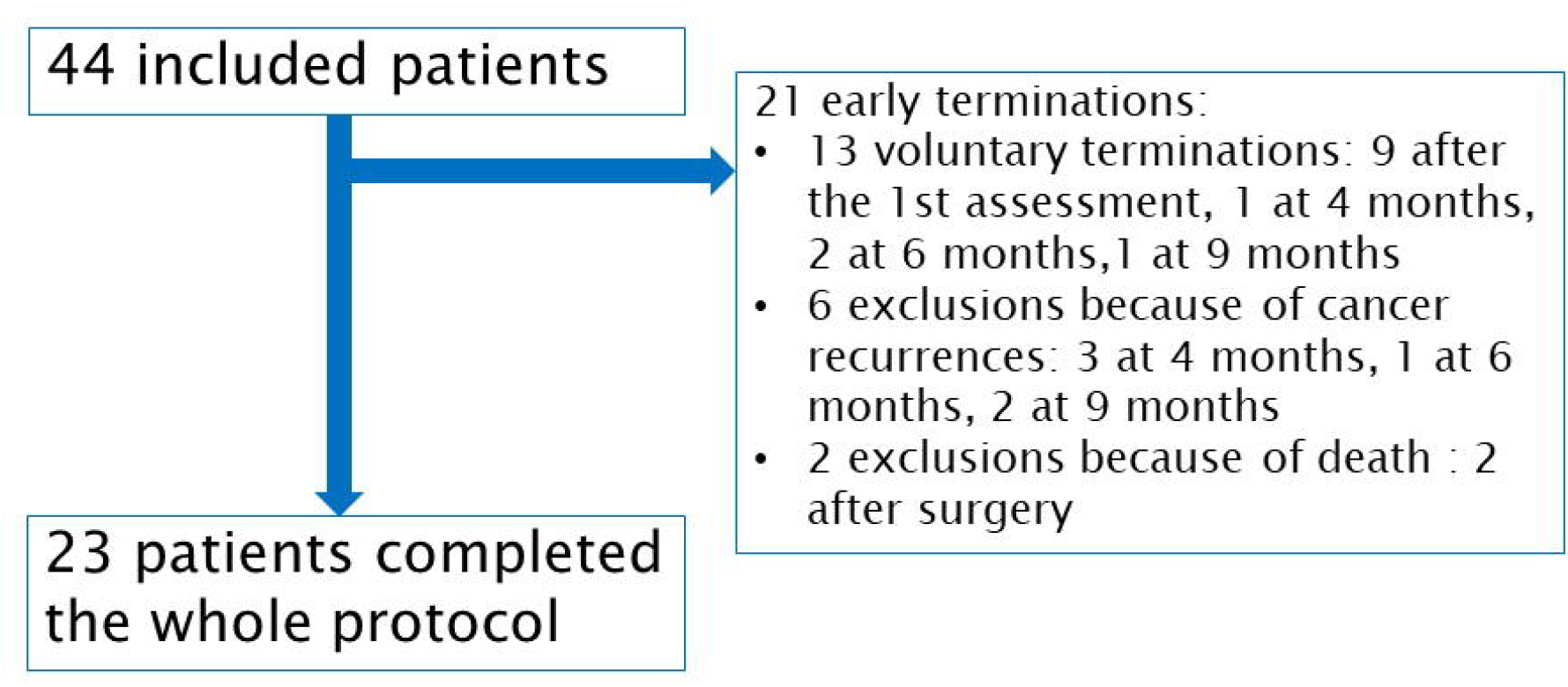
study flowchart

Twelve complained of MT (27.2% of the whole sample; 52.2% of the 23 patients who completed the whole study). MT occurred at different moments (19 occurrences), mainly during the treatment phase (periods 2 to 4). The number and the moment of MT are presented in Figure 2B. There were no significant demographic characteristics differences at inclusion, between MT and no MT status. At Moment 2 (after surgery, if any), two patients complained about MT; at Moment 3 (middle of radiotherapy or radiochemotherapy, if any), four patients; at Moment 4 (end of radiotherapy or radiochemotherapy, if any), eight patients; at Moment 5 (3 months after the end of treatments), four patients; at Moment 6 (six months after the end of treatments), one patient. Figure S1 shows the evolution of the salivary flow according to the different moments and the MT status.

**Figure 2.**
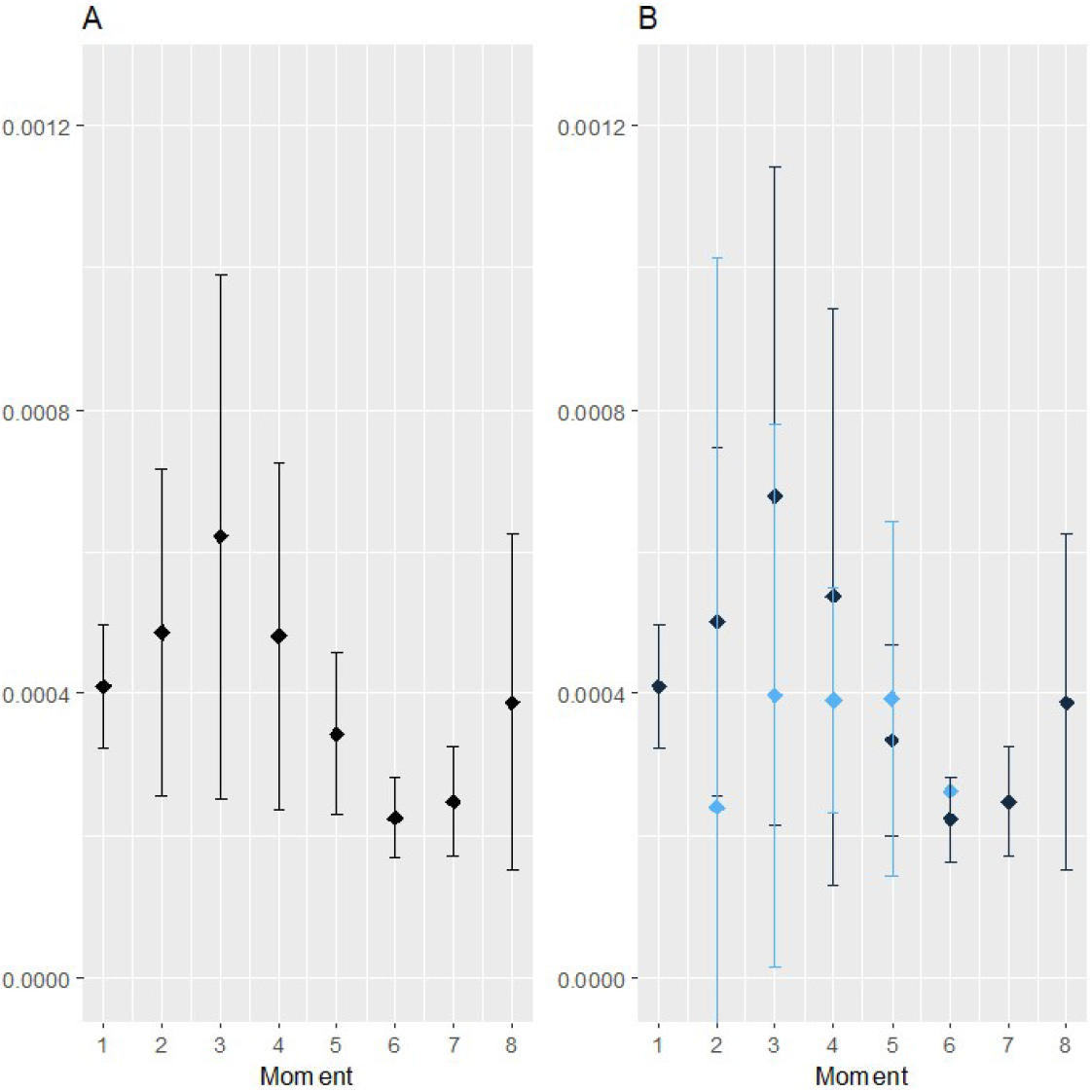
evolution of salivary MDA concentration (µmol/10 min) during the follow-up. A = global results on the whole population. B = according to the metallic taste status (Blue = metallic taste, black = no metallic taste) Moments: (m) moment 1); after surgery if any (m2); in the middle (m3) and at the end (m4) of the radiotherapy if any; at 3 (m5)-6 (m6)-9 (m7) and 12 (m8) months.

### Salivary Malondialdehyde Levels

Figure 2A represents the evolution of salivary MDA concentration according to the different moments. Baseline salivary MDA levels varied widely among participants, with a mean of 0.11 µM (standard deviation [SD] 0.06 µM). Over the one-year follow-up, MDA levels showed a significant increase, peaking at the middle of the radiotherapy (mean 0.26 µM, SD 0.27 µM, p-value of the comparison between Moments 1 and 3=0.034) before declining slightly (at 12 months: mean 0.15 µM, SD 0.1 µM, p=0.125 in comparison with Moment 1). Figure 2B represents the evolution of salivary MDA concentration and the MT status during the one-year-follow-up. Wilcoxon tests revealed no significant link between salivary MDA levels and metallic taste (p=0.44, 0.96, 1, and 0.92 for Moments 2, 3, 4 and 5).

### MT and lactoferrin mouthwash

Figure 3 shows the efficacy of bovine lactoferrin mouthwash in patients complaining about MT. Lactoferrin mouthwash helped to alleviate at least partially MT in 12 out of the 19 MT occurrence (efficacy rate: 63.2%).

**Figure 3.**
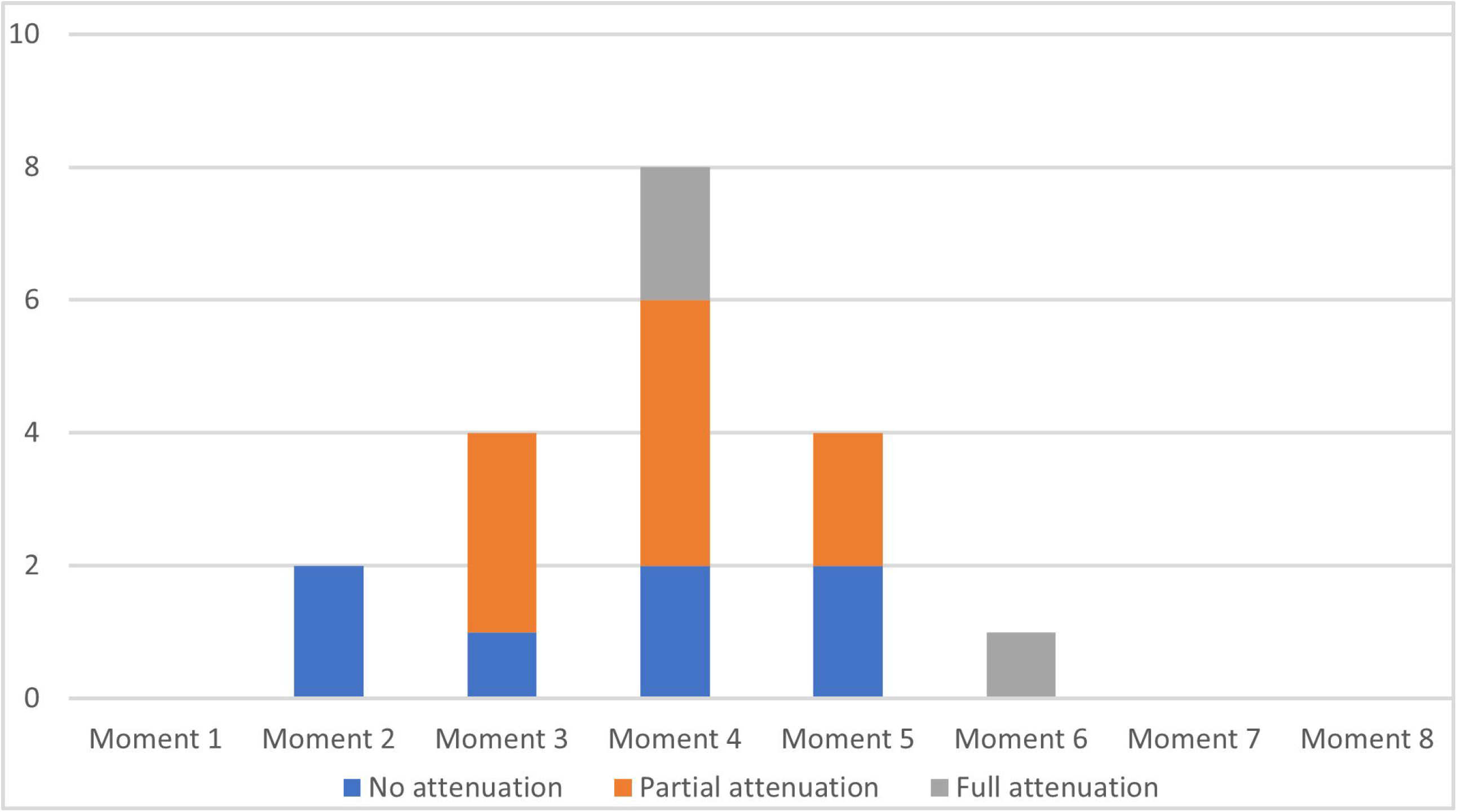
qualitative efficacy of lactoferrin mouthwash in patients with metallic taste according to the evaluation moment, based on patient-reported perceptions of attenuation across three modalities.

### Other salivary analyzes

Baseline salivary protein mean was 0.98 mg/ml (SD 0.73 mg/ml). Over the one-year follow-up, protein levels showed a stable concentration (Figure S2A). Figure S2B represents the evolution of salivary protein concentration over time in two groups (MT vs no-MT). Wilcoxon tests revealed no significant link between salivary protein concentration and MT (all p>0.05).

Baseline salivary antioxidant properties (ORAC) mean was 1663µmol/l (SD 892 µmol/l). Over the one-year follow-up, antioxidant properties remained stable (Figure S3A). Figure S3B represents the evolution of salivary antioxidant properties over time in two groups (MT vs no-MT). Wilcoxon tests revealed no significant link between salivary antioxidant properties and MT (all p>0.05).

Baseline catalase activity mean was 162 IU/ml (SD 148 IU/ml). Over the one-year follow-up, protein levels showed a stable concentration (Figure S4A). Figure S4B represents the evolution of salivary catalase activity over time in two groups (MT vs no-MT). Wilcoxon tests revealed no significant link between salivary protein concentrations and MT (all p>0.05).

Baseline SuperOxide Dismutase (SOD) activity mean was 6.05 IU/ml (SD 5.10 IU/ml). Over the one-year follow-up, SOD showed a stable concentration (Figure S5A). Figure S5B represents the evolution of salivary SOD activity over time in two groups (MT vs no-MT). Wilcoxon tests revealed no significant link between salivary protein levels and MT (all p>0.05).

Baseline salivary GSTp mean was 38.4 ng/ml (SD 41.8 ng/ml). Over the one-year follow-up, GSTp levels slightly decreased after surgery and/or radiotherapy (period 2-4) and returned to baseline later (Figure S6A). Figure S6B represents the evolution of salivary GSTp concentration over time in two groups (MT vs no-MT). Wilcoxon tests revealed no significant link between salivary GSTp concentration and MT (all p>0.05).

## Discussion

Our study aimed to explore the relationship between salivary markers, especially MDA levels, and MT in patients undergoing treatment for HNC.

### MDA levels and MT

The current findings indicate a significant increase in salivary MDA levels during the radiotherapy or radiochemotherapy phase (moments 3 and 4 of Figure 2), which subsequently declined, while other salivary markers such as protein concentration, antioxidant properties, catalase activity, and SOD activity remained relatively stable over the same one-year follow-up (Figure S2-S6).

The observed increase in salivary MDA levels during radiotherapy suggests elevated oxidative stress in patients undergoing cancer treatment. MDA is a well-known marker of lipid peroxidation and oxidative stress, and its elevation during radiotherapy aligns with previous findings that highlight the oxidative damage induced by cancer therapies (7, 13). Agha-Hosseini et al. reported significantly higher salivary MDA levels in patients with oral lichen planus compared to healthy controls, emphasizing the role of oxidative stress in oral pathologies (13). Similarly, Ergun et al. found elevated MDA levels in patients with lichen planus, further supporting the relevance of MDA as a marker for oxidative stress in oral diseases (7). Rai et al. (9) and Saral et al. (16) found similar results. At last, smoking also increases salivary MDA (10, 17). Additionally, as we found, the study by Kaur et al. (8) investigating salivary markers in various oral conditions found significantly elevated MDA levels in patients with 30 oral squamous cell carcinoma and 30 precancerous conditions compared to 30 controls. The subsequent decline in MDA levels post-treatment in our study indicates a reduction in oxidative stress as patients recover. This highlights the role of oxidative stress in HNC and suggests a relationship between MDA levels and symptoms like MT.

However, despite the fluctuations in MDA levels, no significant link with MT was observed, suggesting that factors other than this form of oxidative stress may play a concomitant or more prominent role in the development of MT. Furthermore, despite the 95% confidence intervals largely crossed, the MDA concentration mean was lower in the MT group. Other products of oral lipoperoxidation could lead to the formation of volatile compounds with very low detection thresholds, such as certain aldehydes or ketones (18, 19). Maybe those volatile markers of MT should be looked at instead of MDA, which is a general marker of lipoperoxidation. It should have been possible to evaluate the release of these volatils by measuring nasal effluvia during oral consumption. However such a methodology should be adapted to be implemented to a population of cancer patients. Results on lactoferrin supports the oxidation hypothesis. Indeed, lactoferrin is an iron-binding milk protein, known for its antimicrobial, antiviral, immunostimulating and anti-inflammatory properties (15, 20). It has been proposed as a potential therapeutic agent for managing taste disturbances (6). In our study, the lactoferrin mouthwash helped to alleviate MT in 12 MT occurrences out of 19 (63.2%), which supports the oxidation hypothesis.

Basically, the hypothesis of lipoperoxidation still holds but needs further explorations regarding identification and measurement of the molecules at the origin of MT.

### Other salivary markers and MT

Previous research has shown that salivary protein composition can be altered in various conditions, including cancer therapy, impacting oral health and potentially taste perception (21, 22). Moreover, catalase and SOD are vital in managing oxidative stress, and their activity can be indicative of the overall oxidative status within the oral environment (23, 24). Indeed, our analysis revealed no significant link with MT. These findings suggest that the biochemical changes related to redox status in saliva during cancer treatment do not directly contribute to the development of MT specifically. Indeed those markers had been previously tested with taste disorders including MT (25), without identifying subgroups expressing specific taste disorders associated with specific marker modifications. Moreover, Walliczek-Dworschak et al. compared, among other parameters, 81 patients with taste disorder (the causes were not specified) including 11 patients with MT, to 40 healthy volunteers (26). They showed similar mean salivary protein, catalase concentrations (8.8 (SD10.5) and 10.7 (SD 13.2), p=0.40) and total antioxidant status (476.4 (SD 388.4) and 992.3 (SD 596.5), p=0.09). However, for other taste disorders (agueusia, hypogueusia, bitter taste disorder, salt dysgueusia), authors showed a significantly higher TAC compared to healthy subjects.

### Limitations

A limitation of our study is the small sample size, which may have reduced the statistical power to detect significant associations. Additionally, the reliance on self-reported measures for MT, without incorporating objective taste tests, may limit the robustness of our findings. Future studies should aim to include larger cohorts, comparisons of HNC and healthy volunteers and a combination of subjective and objective measures to provide a more comprehensive understanding of the relationship between salivary markers and MT.

## Conclusion

Our study provides valuable insights into the changes in salivary markers during cancer in the global population, regardless the MT status. The significant increase in salivary MDA levels highlighted the oxidative stress induced by radiotherapy. However there was no significant difference in the mean salivary MDA levels between patients reporting MT and those who did not. As MDA is the final stable product of lipoperoxidation, it doesn’t seem the lipoperoxidation could explain MT. However the palliation effect of lactoferrin mouthwash suggests potential benefits of antioxidants. Is there another marker that would stimulate more the retronasal path than taste which could trigger MT? Is the oral microbiota involved? Overall, our findings underscore the need for larger, more comprehensive studies focused on HNC patients treated by radiotherapy and or radiochemotherapy to fully elucidate the mechanisms underlying MT and to develop effective interventions for managing this distressing symptom in cancer patients.

## Data Availability

All data produced are available online at https://hal.science/hal-04750951

