## Supplements for "Exploring the Role of Oxidative Stress in Metallic Taste During Head and Neck Cancer Treatment: A Study of Salivary Malondialdehyde and Therapeutic Interventions"

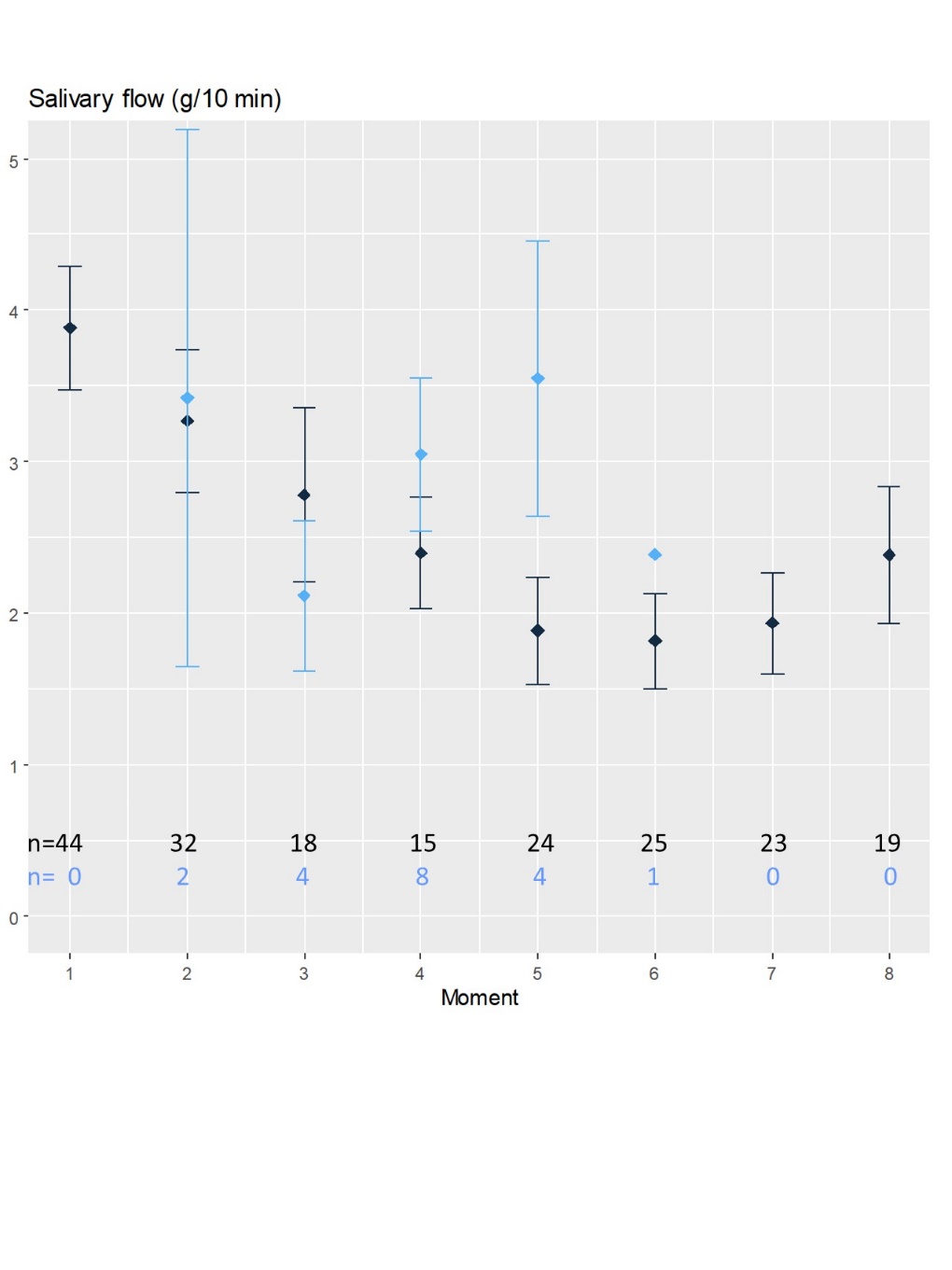

Figure S1: evolution of the salivary flow according to the different moments and the MT status.

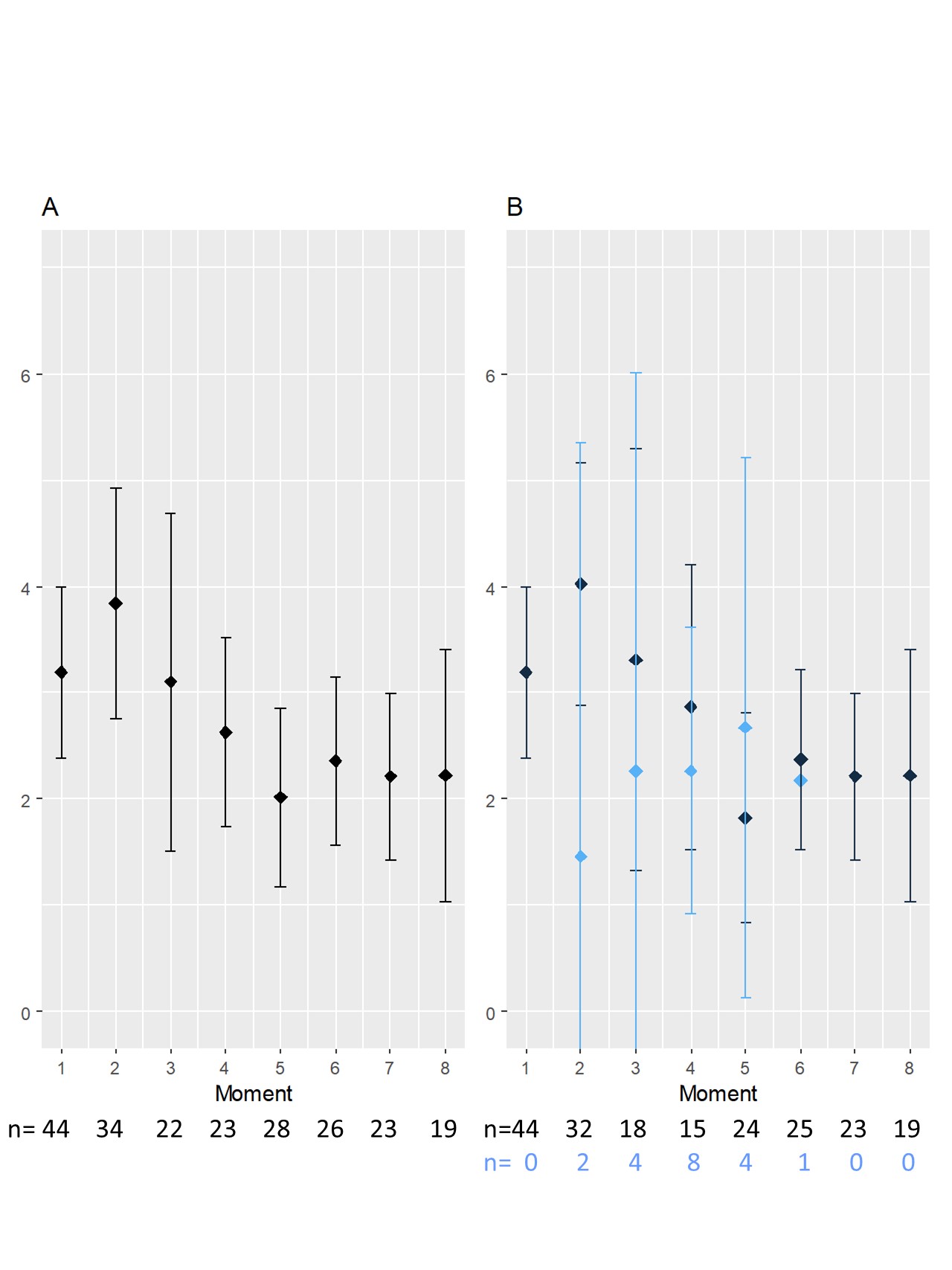

Figure S2: evolution of salivary protein concentration

A: global results of protein concentration (mg/ml) over time. B = protein concentration over time in two groups (Blue = metallic taste, black = no metallic taste)

Moments: (m) moment 1); after surgery if any (m2); in the middle (m3) and at the end (m4) of the radiotherapy if any; at 3 (m5)-6 (m6)-9 (m7) and 12 (m8) months.

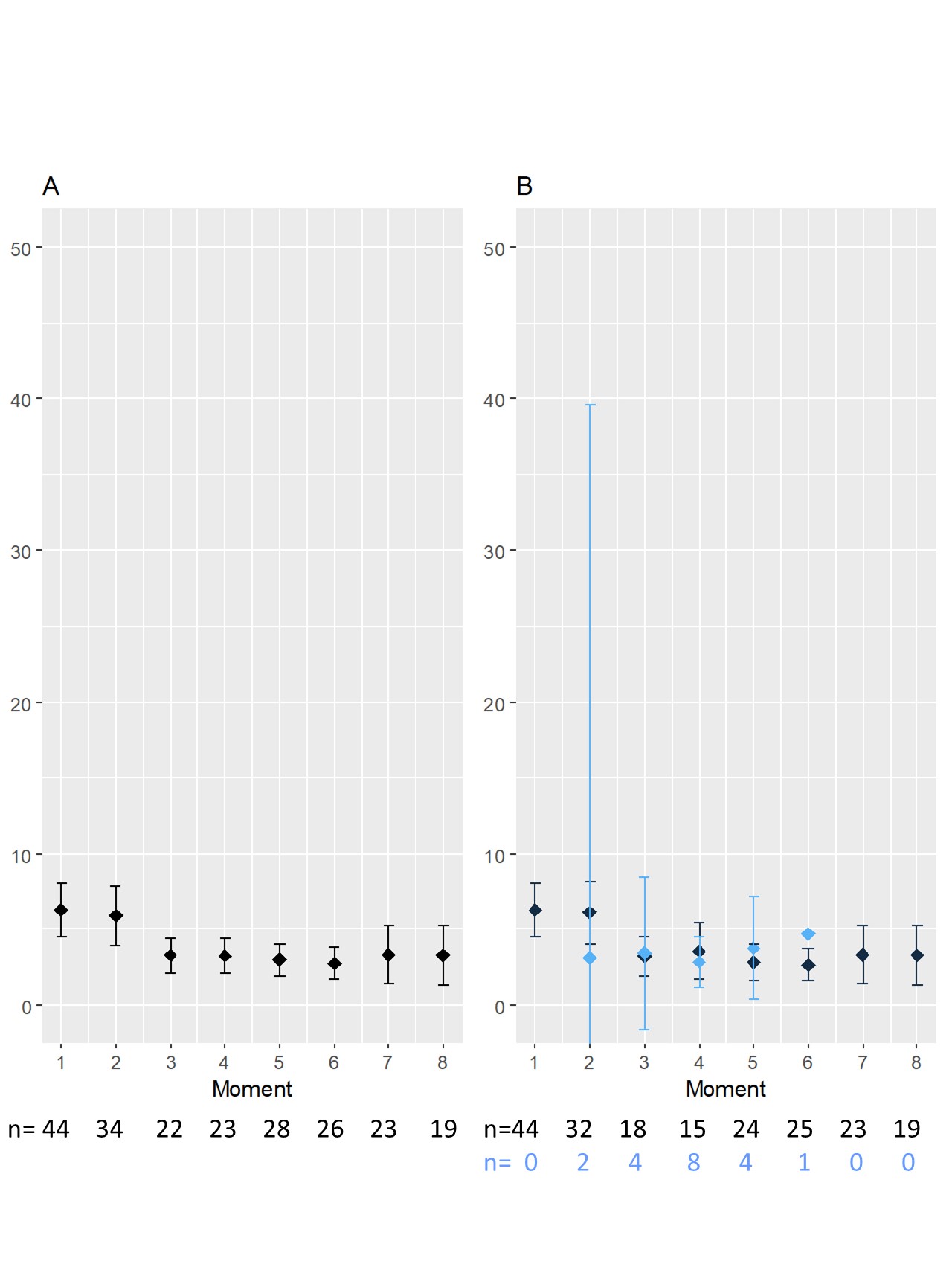

Figure S3: salivary antioxidant properties

A: global results of salivary antioxidant properties (µmol/ 10 min) over time. B = salivary antioxidant properties (µmol/l) over time in two groups (Blue = metallic taste, black = no metallic taste)

Moments: (m) moment 1); after surgery if any (m2); in the middle (m3) and at the end (m4) of the radiotherapy if any; at 3 (m5)-6 (m6)-9 (m7) and 12 (m8) months.

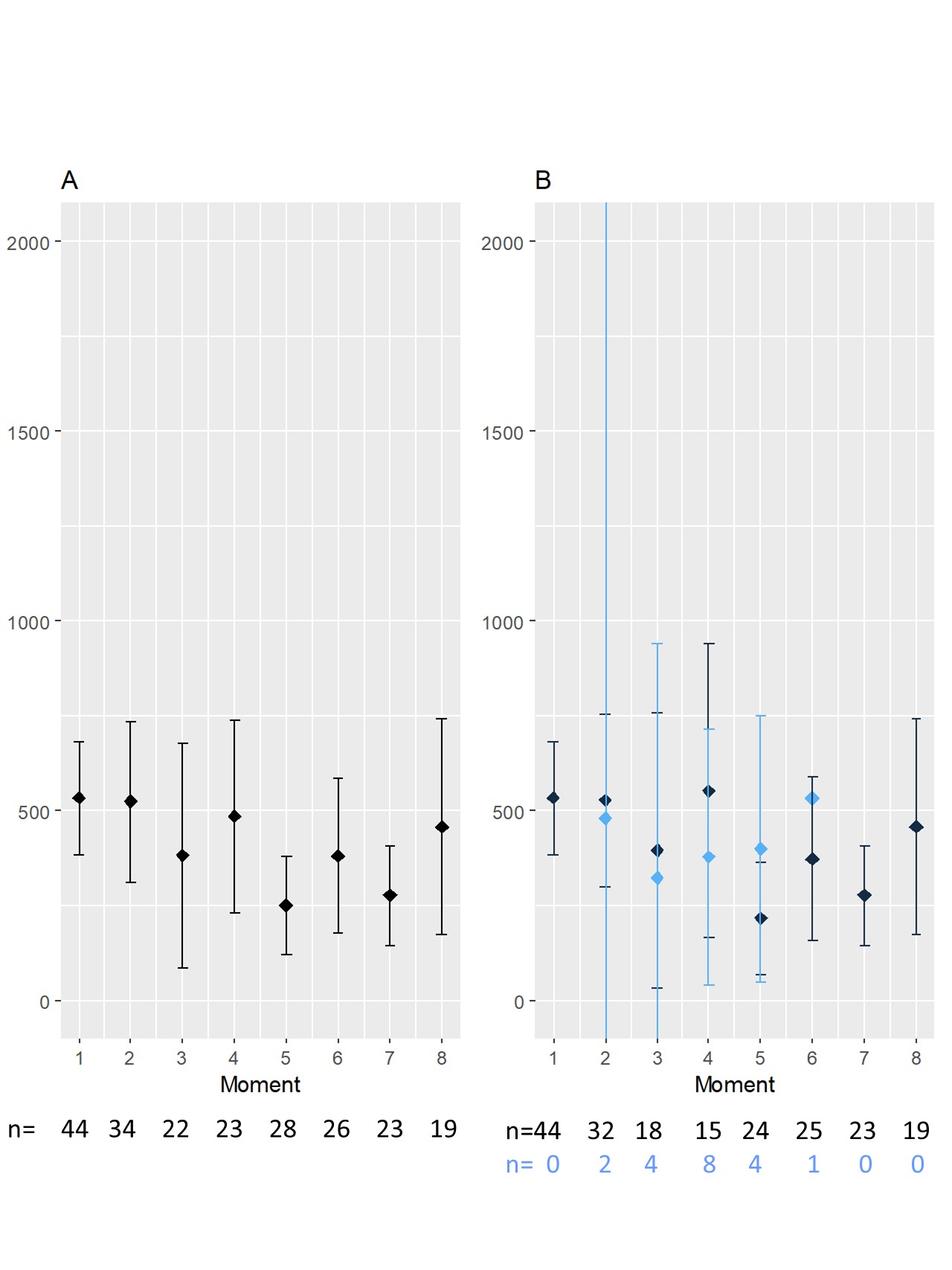

Figure S4: salivary catalase concentration

A: global results of salivary catalase concentration (IU/10 min) over time. B = salivary catalase concentration (IU/ml) over time in two groups (Blue = metallic taste, black = no metallic taste)

Moments: (m) moment 1); after surgery if any (m2); in the middle (m3) and at the end (m4) of the radiotherapy if any; at 3 (m5)-6 (m6)-9 (m7) and 12 (m8) months.

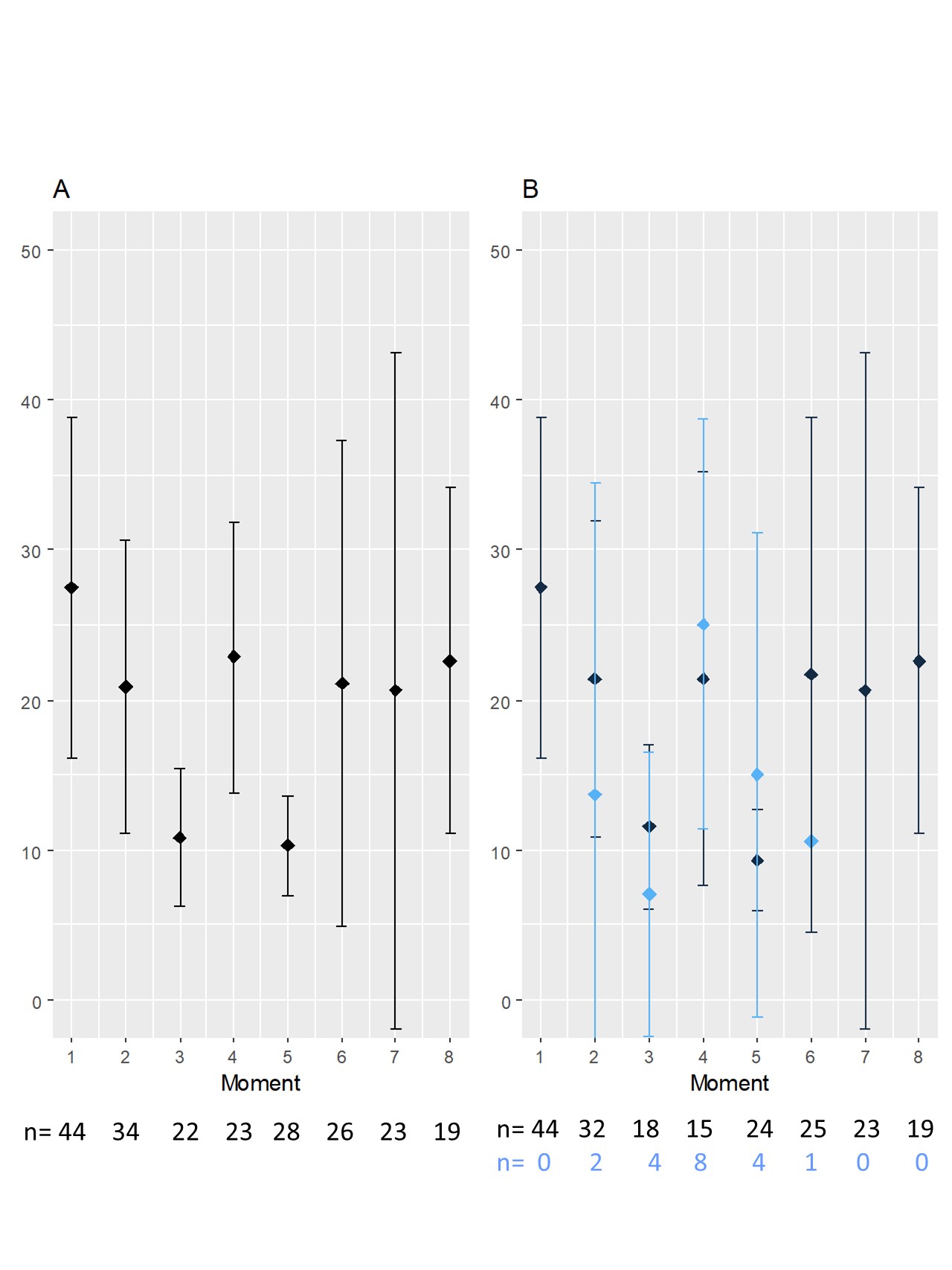

Figure S5: salivary superoxide dismutase concentration

A: global results of salivary superoxide dismutase concentration (IU/ml) over time. B = salivary superoxide dismutase concentration (IU/10 min) over time in two groups (Blue = metallic taste, black = no metallic taste)

Moments: (m) moment 1); after surgery if any (m2); in the middle (m3) and at the end (m4) of the radiotherapy if any; at 3 (m5)-6 (m6)-9 (m7) and 12 (m8) months.

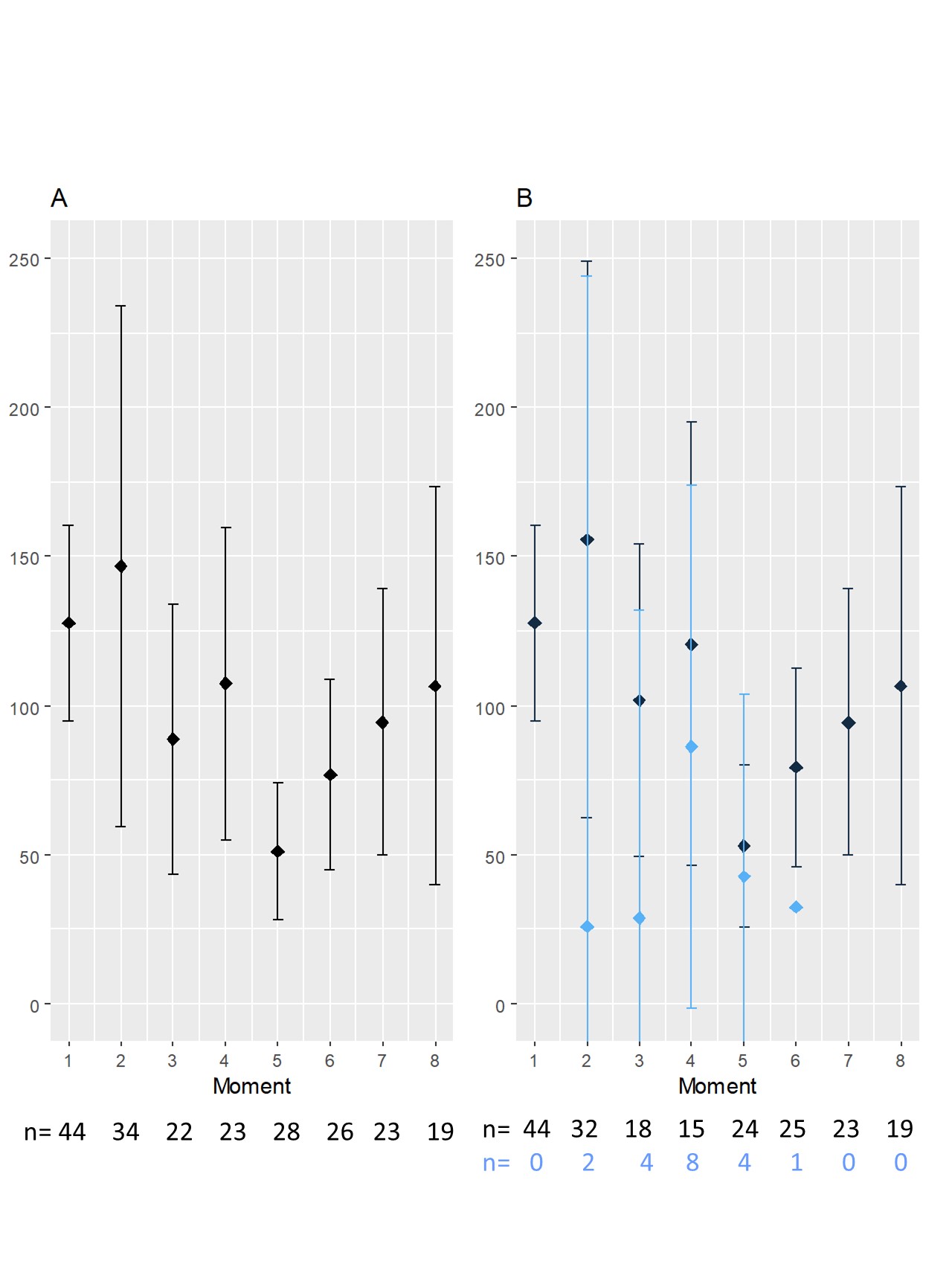

Figure S6: salivary GSTP1 concentration

A: global results of salivary GSTP1 concentration (IU/10min) over time. B = salivary GSTP1 concentration (IU/ml) over time in two groups (Blue = metallic taste, black = no metallic taste)

Moments: (m) moment 1); after surgery if any (m2); in the middle (m3) and at the end (m4) of the radiotherapy if any; at 3 (m5)-6 (m6)-9 (m7) and 12 (m8) months.
